# Artificial Intelligence for COVID-19 Risk Classification in Kidney Disease: Can Technology Unmask an Unseen Disease?

**DOI:** 10.1101/2020.06.15.20131680

**Authors:** Caitlin Monaghan, John W. Larkin, Sheetal Chaudhuri, Hao Han, Yue Jiao, Kristine M. Bermudez, Eric D. Weinhandl, Ines A. Dahne-Steuber, Kathleen Belmonte, Luca Neri, Peter Kotanko, Jeroen P. Kooman, Jeffrey L. Hymes, Robert J. Kossmann, Len A. Usvyat, Franklin W. Maddux

## Abstract

**Background:** We developed two unique machine learning (ML) models that predict risk of: 1) a major COVID-19 outbreak in the service county of a local HD population within following week, and 2) a hemodialysis (HD) patient having an undetected SARS-CoV-2 infection that is identified after following 3 or more days.

**Methods:** We used county-level data from United States population (March 2020) and HD patient data from a network of clinics (February-May 2020) to develop two ML models. First was a county-level model that used data from general and HD populations (21 variables); outcome of a COVID-19 outbreak in a dialysis service area was defined as a clinic being located in one of the national counties with the highest growth in COVID-19 positive cases (number and people per million (ppm)) in general population during 22-28 Mar 2020. Second was a patient-level model that used HD patient data (82 variables) to predict an individual having an undetected SARS-CoV-2 infection that is identified in subsequent ≥3 days.

**Results:** Among 1682 counties with dialysis clinics, 82 (4.9%) had a COVID-19 outbreak during 22-28 Mar 2020. Area under the receiver operating characteristic curve (AUROC) for the county-level model was 0.86 in testing dataset. Top predictor of a county experiencing an outbreak was the COVID-19 positive ppm in the general population in the prior week. In a select group (n=11,664) used to build the patient-level model, 28% of patients had COVID-19; prevalence was by design 10% in the testing dataset. AUROC for the patient-level model was 0.71 in the testing dataset. Top predictor of an HD patient having a SARS-CoV-2 infection was mean pre-HD body temperature in the prior week.

**Conclusions:** Developed ML models appear suitable for predicting counties at risk of a COVID-19 outbreak and HD patients at risk of having an undetected SARS-CoV-2 infection.

## Introduction

The 2019 coronavirus disease (COVID-19) pandemic is challenging the world’s healthcare systems, including bringing complexities to maintenance of dialysis in people with end stage kidney disease (ESKD) (1-5). In the United States, most ESKD patients are treated by outpatient hemodialysis (HD) where social distancing can be difficult and heightened infection control measures are required (e.g. temperature screenings, double-masking, isolation treatments/shifts/clinics) (1-5). ESKD patients are typically older and have multiple comorbidities, placing the population at higher risks for requiring intensive care and dying if affected by COVID-19 (6-12).

Early reports from the United States show an 11% COVID-19 mortality in ESKD (13), which is higher than the 3.2% COVID-19 mortality in the national population (14). This is not unexpected with reports from China and Europe suggesting a 16% and 23% COVID-19 mortality in ESKD (15, 16). Albeit the high mortality rate, an impaired immune response may render dialysis patients more frequently asymptomatic when infected by SARS-CoV-2 (15, 16). In both the general and EKSD populations, the most prevalent symptoms of COVID-19 at presentation are fever (11%-66% dialysis & 82% general population) and cough (57% dialysis & 62% general population) (15, 17, 18). The less frequent occurrence of signs and symptoms indicative of COVID-19 in dialysis patients could be making the outbreak even more challenging to manage.

Dialysis providers routinely capture patient/clinical data during care. The robust data collected during HD treatments (generally thrice weekly) provide unique opportunities to leverage artificial intelligence (AI) in predicting COVID-19 outcomes. AI modeling helped identify onset of the outbreak in China (19, 20) and is currently being used to help with early detection of areas/individuals in the general population at risk of being affected by COVID-19 (21-23).

As part of an healthcare operations effort in response to the COVID-19 outbreak, an integrated kidney disease healthcare company aimed to develop two unique machine learning (ML) prediction models that identify risk of: 1) a major COVID-19 outbreak in a dialysis service county, and 2) a HD patient having an undetected severe acute respiratory syndrome coronavirus-2 (SARS-CoV-2) infection. We analyzed the performance of these models to determine their possible utility for testing in the HD population.

## Materials and Methods

### General

An integrated kidney disease healthcare company (Fresenius Medical Care, Waltham, MA, United States) used COVID-19 data on the general United States population and HD patient data from a national network of dialysis clinics (Fresenius Kidney Care, Waltham, MA, United States) to develop two unique ML models that predict risk of: 1) a major COVID-19 outbreak in the county of dialysis clinic(s) servicing a local HD population in the following week (7 days), and 2) an adult HD patient having an undetected SARS-CoV-2 infection that is identified after the following ≥3 days.

This analysis was performed in adherence with the Declaration of Helsinki under a protocol reviewed by New England Independent Review Board (NEIRB). This retrospective analysis was determined to be exempt and did not require consent (Needham Heights, MA, United States; NEIRB#1-17-1302368-1).

### Populations and Outcomes

#### County Population and COVID-19 Outbreak

For construction of a model to predict a major COVID-19 outbreak in the county for a local HD population, we obtained data on daily COVID-19 positive (COVID-19+) cases in each United States county during 14-28 Mar 2020 from The New York Times COVID-19 dataset (The New York Times Company, New York City, United States) (24). We also captured data on the total United States county population (25). We used retrospective county-level data from local HD populations at the national network of clinics from 08-28 Mar 2020.

We used data from all counties with ≥1 dialysis clinic and >5 HD patients. The outcome of a major COVID-19 outbreak in a dialysis service area (county) was defined was defined by a clinic being located in one of the top 300 national counties that had a combination of the highest (a) growth rate in COVID-19+ cases (number and people per million (ppm)) in the general population, and (b) highest number of COVID-19+ cases ppm in the last week of the observation period (22-28 Mar 2020).

#### Patient Population and SARS-CoV-2 Infection

For the construction of a model to predict individuals with an undetected SARS-CoV-2 infection, we considered data from adult (age ≥18 years) HD patients treated in the national network. Positive arm included data from patients who had ≥1 laboratory (rRT-PCR) confirmed COVID-19+ test as of the end of the observation period (27 Feb 2020 through 04 May 2020). Negative arm included data from patients who: 1) resided in counties with no reported COVID-19 cases as of 12 April 2020, and/or 2) had been laboratory tested and were found COVID-19 negative.

We defined the index date of a HD patient having a SARS-CoV-2 infection as the date of the first recorded suspicion that led to a COVID-19+ test, or the date of the COVID-19+ test in patients without an earlier recorded suspicion. In control patients with a negative COVID-19 test result, the test date was used as the index date. In controls without a test (patients living in counties with no reported cases), the index date was randomly sampled from the positive cases’ index dates occurring before 30 March 2020. This cutoff was chosen to minimize the possibility that control patients were infected but not tested (or reported) until after the date defining negative counties of 12 April 2020. We included data from patients with 1) ≥1 laboratory (of any kind) collected both 1-14 days and 31-60 days before the individual’s prediction date (3 days prior to index date, further defined below), and 2) ≥1 HD treatment both 1-7 days and 31-60 days preceding the prediction date. We excluded data from patients suspected to have COVID-19 and/or pending laboratory testing.

### AI Model Development

#### Software

We used Python version 3.7.7 (Python Software Foundation, Delaware, United States) to build two ML models utilizing the XGBoost package (26).

#### County-Level COVID-19 Outbreak Prediction Model

In the county-level model, we considered county-wide data in the general population for 4 variables and county-wide data on each local HD population based on the county of the dialysis clinic(s) for 17 *a priori* selected variables detailed in **Table 1** and **Figure 1A**. The ML model was trained, validated, and tested using a random 60:20:20% split of the dataset.

**Table 1:** Characteristics of Dialysis Service Areas (Counties) with and without a Major COVID-19 Outbreak in the Subsequent 7 Days

| Variable | Counties without Major Outbreak<br>N (%) or Mean±SD | Counties with Major COVID-19 Outbreak<br>N (%) or Mean±SD |
| --- | --- | --- |
| **County Characteristics of General Population** |  |  |
| Number of Counties | 1600 | 82 |
| COVID-19+ ppm in Prior Week† | 23 | 87 |
| Deaths per COVID-19+ Case in Prior Week† | 0.016884 | 0.021824 |
| Change in COVID-19+ ppm in Prior 2 Weeks† | 5.90 | 15.43 |
| Population Density (Per Square Mile)† | 355 | 1196 |
| **County Characteristics of Local HD Population** |  |  |
| Number of HD Patients | 158070 | 20531 |
| Residing in Assisted Living Facility† | 10% | 11% |
| HD Nonadherence in the Prior 2 Weeks† | 5.2% | 4.9% |
| % Treatments with Pre/Post-HD Body Temperature(s) >100°F in Prior 2 Weeks† | 0.79% | 0.83% |
| Change in 2 Week Mean Number of HD Treatments in Prior 4 Weeks† | 0.9% | 1.1% |
| Change in 2 Week Mean % HD Nonadherence in Prior 4 Weeks† | -0.8% | -0.4% |
| Change in 2 Week Mean % Treatments with Body Temperature(s) >100°F in Prior 4 Weeks† | 0.3% | 0.4% |
| Change in 2 Week Mean HD Treatment Time in Prior 4 Weeks† | 0.04% | 0.01% |
| Change in 2 Week Mean Pre-HD DBP in Prior 4 Weeks† | -0.12% | -0.14% |
| Change in 2 Week Mean Pre-HD Body Temperature in Prior 4 Weeks† | 0.07% | 0.10% |
| Change in 2 Week Mean Pre-HD Weight in Prior 4 Weeks† | 0.01% | -0.13% |
| Change in 2 Week Mean IDWG in Prior 4 Weeks† | 0.9% | 1.2% |
| Change in 2 Week Mean LDH in Prior 4 Weeks† | 4.16% | 1.31% |
| Change in 2 Week Mean Platelets in Prior 4 Weeks† | 0.76% | -1.19% |
| Change in 2 Week Mean Lymphocytes in Prior 4 Weeks† | 5.08% | 4.05% |
| Change in 2 Week Mean Neutrophils in Prior 4 Weeks † | -0.32% | -0.41% |
| Change in 2 Week Mean Monocytes in Prior 4 Weeks† | 2.18% | 3.97% |
| Change in 2 Week Mean Hgb in Prior 4 Weeks† | 0.12% | 0.39% |
| † County-wide variables included in the ML prediction model to classify the 7-day risk of a COVID-19 outbreak in a county with dialysis clinics servicing a local HD population.<br>HD: hemodialysis; CHF: congestive heart failure; BMI: body mass index; IDWG: interdialytic weight gain; Hgb: hemoglobin; N: patient count; SD: standard deviation. 1 mile = 1.6 kilometer. $(100^{\circ}\text{F} - 32) \times 5/9 = 37.8^{\circ}\text{C}$. | | |

**Figure 1:**
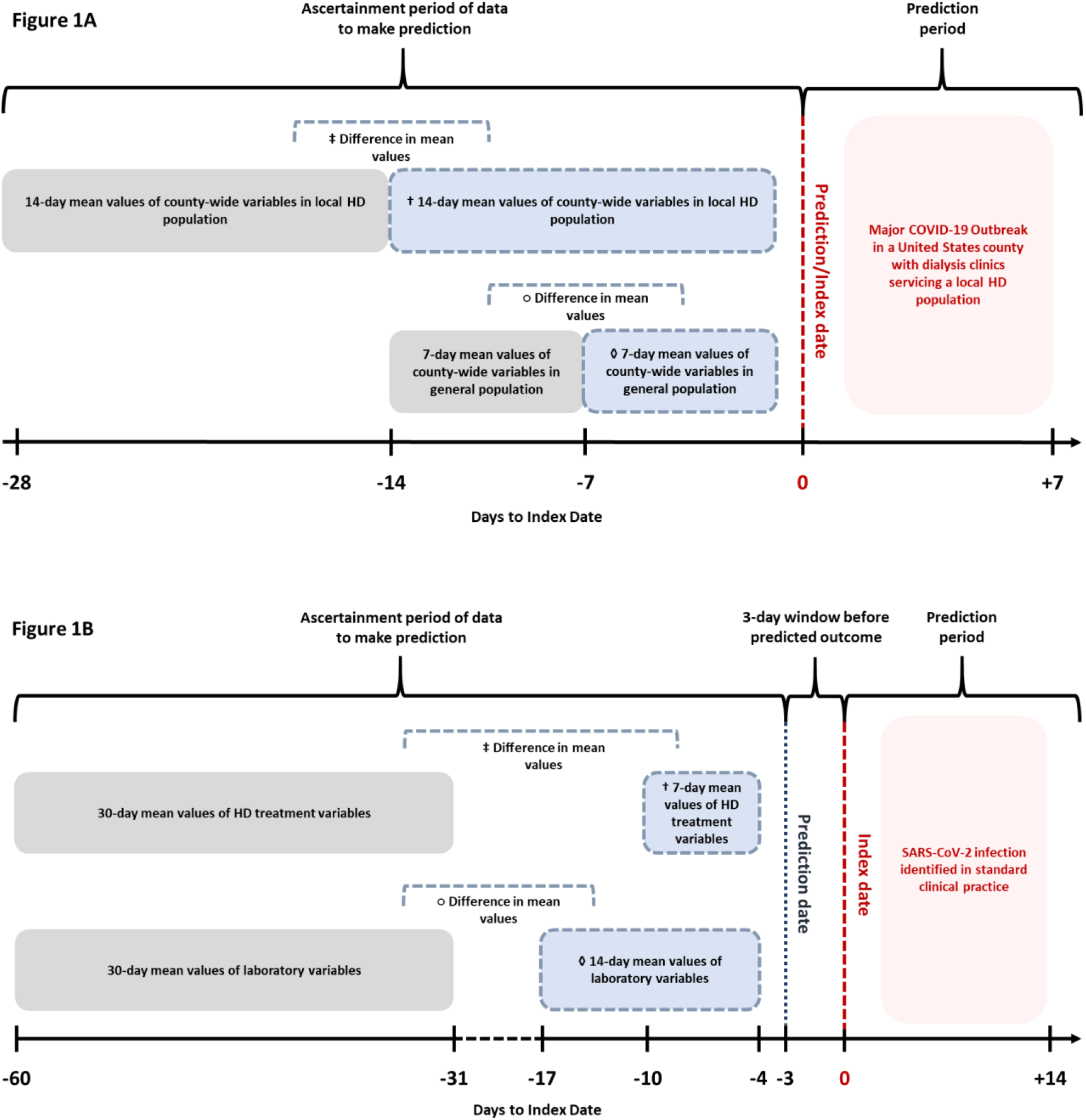
Prediction timelines for the unique county- and patient-level ML models. **A)** Timeline of data ascertainment and prediction of dialysis service areas (counties) with and without a major COVID-19 outbreak in the subsequent 7 days. ML model used county-wide variables in local HD population († mean values 14 days before the index date; ‡ difference in mean values 15-28 days to 1-14 days before the index date) and county-wide variables in general population (◊ 1-7 days before the index date; ○ difference in mean values 8-14 days to 1-7 days before the index date) for prediction of major COVID-19 outbreak in a United States county. **B)** Timeline of data ascertainment and prediction of HD patients with and without SARS-CoV-2 infection identified in the subsequent ≥3 days. ML model used HD treatment variables († mean values 1-7 days before the prediction date; ‡ difference in mean values 31-60 days to 1-7 days before the prediction date) and laboratory variables (◊ mean values 1-14 days before the prediction date; ○ difference in mean values 31-60 days to 1-14 days before the prediction date) for prediction of SARS-CoV-2 infection.

#### Patient-Level SARS-CoV-2 Infection Prediction Model

In the patient-level model, we used 82 *a priori* selected treatment/laboratory variables (**Tables 3 and 4; Figure 1B**) up to the individually defined prediction date (3 days prior to the index date defined above) to predict the risk of a SARS-CoV-2 infection being identified in the following ≥3 days. This is intended to yield individual predictions at least 3 days in advance of symptoms that warranted testing. We used a 60:20:20% randomized split of COVID-19+ samples for the training, validation, and testing datasets, and added the same number of COVID-19 negative patients to only the training and validation datasets. The testing dataset used to evaluate final model performance had a higher number of COVID-19 negative samples added to more closely match the prevalence observed in the overall national HD population.

**Table 3:** Clinical and Treatment Characteristics of HD Patients with and without SARS-CoV-2 Infection Identified in the Subsequent ≥3 Days

| Variable | Unaffected Patients<br>N (%) or Mean±SD | COVID-19+ Patients<br>N (%) or Mean±SD |
| --- | --- | --- |
| Pre-HD Sitting SBP (mmHg)<sup>†</sup> | 8415, 149.73±23.35 | 3249, 145.95±23.38 |
| Change in Pre-HD Sitting SBP (mmHg)<sup>‡</sup> | 8415, 0.07±16.45 | 3249, -0.95±16.72 |
| Pre-HD Sitting DBP (mmHg)<sup>†</sup> | 8415, 77.60±14.32 | 3249, 75.44±13.30 |
| Change in Pre-HD Sitting DBP (mmHg)<sup>‡</sup> | 8415, -0.03±9.44 | 3249, -0.56±9.15 |
| Pre-HD Weight (kg)<sup>†</sup> | 8415, 86.80±24.09 | 3249, 85.11±25.32 |
| Change in Pre-HD Weight (kg)<sup>‡</sup> | 8415, -0.20±2.50 | 3249, -0.75±2.80 |
| Pre-HD Body Temperature (°F)<sup>†</sup> | 8415, 97.50±0.63 | 3249, 97.73±0.67 |
| Change in Pre-HD Body Temperature (°F)<sup>‡</sup> | 8415, 0.09±0.59 | 3249, 0.28±0.66 |
| Post-HD Sitting SBP (mmHg)<sup>†</sup> | 8415, 141.20±22.07 | 3249, 140.75±21.17 |
| Change in Post-HD Sitting SBP (mmHg)<sup>‡</sup> | 8415, 1.39±15.63 | 3249, 2.13±15.52 |
| Post-HD Sitting DBP (mmHg)<sup>†</sup> | 8415, 73.91±12.90 | 3249, 73.41±12.24 |
| Change in Post-HD Sitting DBP (mmHg)<sup>‡</sup> | 8415, 0.80±8.83 | 3249, 0.99±8.78 |
| Post-HD Body Temperature (°F)<sup>†</sup> | 8415, 97.60±0.57 | 3249, 97.73±0.62 |
| Change in Post-HD Body Temperature (°F)<sup>‡</sup> | 8415, 0.07±0.50 | 3249, 0.20±0.58 |
| Pre-HD Respirations per Minute<sup>†</sup> | 8415, 17.73±1.25 | 3249, 17.74±1.09 |
| Change in Pre-HD Respirations per Minute<sup>‡</sup> | 8415, 0.01±1.02 | 3249, 0.02±1.01 |
| Pre-HD Pulse (BPM)<sup>†</sup> | 8415, 79.09±12.28 | 3249, 78.60±11.90 |
| Change in Pre-HD Pulse (BPM)<sup>‡</sup> | 8415, 0.23±7.49 | 3249, 1.06±7.55 |
| Post-HD Respirations per Minute<sup>†</sup> | 8415, 17.61±1.15 | 3249, 17.69±1.10 |
| Change in Post-HD Respirations per Minute<sup>‡</sup> | 8415, 0.002±0.95 | 3249, 0.01±0.99 |
| Post-HD Pulse (BPM)<sup>†</sup> | 8415, 76.87±11.44 | 3249, 77.42±11.13 |
| Change in Post-HD Pulse (BPM)<sup>‡</sup> | 8415, -0.32±7.45 | 3249, 0.99±7.81 |
| IDWG (kg)<sup>†</sup> | 8343, 2.23±1.28 | 3217, 1.90±1.23 |
| Change in IDWG (kg) <sup>‡</sup> | 8321, 0.02±1.01 | 3201, -0.22±1.04 |
| Post-HD Weight Loss (kg)<sup>†</sup> | 8410, -2.26±1.12 | 3249, -2.02±1.04 |
| Change in Post-HD Weight Loss (kg)<sup>‡</sup> | 8409, -0.03±0.75 | 3248, 0.13±0.75 |
| Post-HD Body Temperature Change<sup>†</sup> | 8415, 0.11±0.65 | 3249, 0.001±0.69 |
| Change in Post-HD Body Temperature Change<sup>‡</sup> | 8415, -0.02±0.66 | 3249, -0.08±0.69 |
| Post-HD Respirations per Minute Change<sup>†</sup> | 8415, -0.12±1.01 | 3249, -0.05±0.92 |
| Change in Post-HD Respirations per Minute Change<sup>‡</sup> | 8415, -0.01±1.07 | 3249, -0.01±1.03 |
| Post-HD Pulse Change (BPM)<sup>†</sup> | 8415, -2.21±8.77 | 3249, -1.18±8.49 |
| Change in Post-HD Pulse Change (BPM) <sup>‡</sup> | 8415, -0.54±7.85 | 3249, -0.08±4.86 |
| % HD Treatments with Nasal Oxygen Administered<sup>†</sup> | 8415, 0.06±0.20 | 3249, 0.06±0.18 |
| Change in % HD Treatments with Nasal Oxygen Administered<sup>‡</sup> | 8415, 0.006±0.15 | 3249, 0.007±0.14 |

All variables were included in the ML prediction model to classify the risk of an individual HD patient having a SARS-CoV-2 infection being identified in the following $\geq 3$ days.
<sup>†</sup> Mean values of HD treatment variables 1-7 days before the prediction date (i.e. 3 days before suspicion of SARS-CoV-2 infection in standard clinical practice).
<sup>‡</sup> Mean values of the difference in HD treatment variables 31-60 days to 1-7 days before the prediction date.
HD: hemodialysis; SBP: systolic blood pressure; DBP: diastolic blood pressure; IDWG: interdialytic weight gain; Post-HD Weight Loss: post-HD minus pre-HD weight (kg); N: patient count; SD: standard deviation. (100°F – 32) × 5/9 = 37.8°C

**Table 4:** Laboratory Characteristics of HD Patients with and without SARS-CoV-2 Infection Identified in the Subsequent ≥3 Days

| Variable | Unaffected Patients<br>N (%) or Mean±SD | COVID-19+ Patients<br>N (%) or Mean±SD |
| --- | --- | --- |
| Albumin (g/dL)<sup>†</sup> | 4995, 3.7±0.4 | 1486, 3.7±0.5 |
| Change in Albumin (g/dL)<sup>‡</sup> | 4655, -0.01±0.25 | 1432, -0.02±0.3 |
| Creatinine (mg/dL)<sup>†</sup> | 4968, 7.9±2.9 | 1487, 8.2±3.0 |
| Change in Creatinine (mg/dL)<sup>‡</sup> | 4615, 0.04±1.4 | 1423, 0.1±1.4 |
| Bicarbonate (mmol/L)<sup>†</sup> | 4991, 24.2±3.1 | 1496, 24.3±3.1 |
| Change in Bicarbonate (mmol/L)<sup>‡</sup> | 4613, -0.1±3.1 | 1435, -0.2±3.0 |
| BUN (mg/dL)<sup>†</sup> | 5097, 53.8±18.1 | 1654, 55.1±18.5 |
| Change in BUN (mg/dL)<sup>‡</sup> | 4835, -0.1±16.2 | 1614, -0.3±16.1 |
| URR<sup>†</sup> | 4852, 74.9±6.4 | 1569, 74.9±6.1 |
| Change in URR<sup>‡</sup> | 4511, 0.06±5.9 | 1507, 0.05±5.7 |
| Sodium (mmol/L)<sup>†</sup> | 4855, 137.5±3.4 | 1465, 137.3±3.5 |
| Change in Sodium (mmol/L)<sup>‡</sup> | 4488, -0.07±2.9 | 1404, -0.4±3.2 |
| Potassium (mmol/L)<sup>†</sup> | 5595, 4.8±0.7 | 1790, 4.8±0.7 |
| Change in Potassium (mmol/L)<sup>‡</sup> | 5278, 0.01±0.6 | 1746, 0.01±0.6 |
| Phosphate (mg/dL)<sup>†</sup> | 5357, 5.5±1.8 | 1686, 5.2±1.7 |
| Change in Phosphate (mg/dL)<sup>‡</sup> | 5054, 0.02±1.5 | 1646, -0.02±1.4 |
| Calcium (mg/dL)<sup>†</sup> | 5397, 8.8±0.7 | 1725, 8.9±0.7 |
| Change in Calcium (mg/dL)<sup>‡</sup> | 5100, -0.003±0.6 | 1680, -0.02±0.6 |
| Corrected Calcium (mg/dL)<sup>†</sup> | 4783, 9.0±0.7 | 1410, 9.1±0.7 |
| Change in Corrected Calcium (mg/dL)<sup>‡</sup> | 4365, 0.007±0.5 | 1347, 0.005±0.6 |
| iPTH (pg/mL)<sup>†</sup> | 3428, 481.3±423.6 | 1178, 494.8±484.6 |
| Change in iPTH (pg/mL)<sup>‡</sup> | 2445, -26.4±303.8 | 855, -30.2±294.5 |
| Ferritin (ng/mL)<sup>†</sup> | 2827, 1036.8±553.9 | 1020, 1213.5±981.1 |
| Change in Ferritin (ng/mL)<sup>‡</sup> | 1509, 29.7±516.4 | 540, 166.9±761.1 |
| TSAT (%)<sup>†</sup> | 4870, 32.1±14.2 | 1446, 30.3±14.4 |
| Change in TSAT (%)<sup>‡</sup> | 4460, -0.1±15.1 | 1375, -2.0±15.7 |
| Hgb (g/dL)<sup>†</sup> | 8415, 10.6±1.3 | 3249, 10.6±1.3 |
| Change in Hgb (g/dL)<sup>‡</sup> | 8415, 0.03±1.1 | 3249, 0.1±1.2 |
| Platelet Count (x 10<sup>9</sup>/L)<sup>†</sup> | 3891, 199.6±75.0 | 1298, 195.2±79.4 |
| Change in Platelet Count (x 10<sup>9</sup>/L)<sup>‡</sup> | 3501, -1.3±52.3 | 1208, -9.5±56.9 |
| WBC Count (x 10<sup>9</sup>/L)<sup>†</sup> | 4696, 7.1±2.5 | 1463, 6.6±2.4 |
| Change in WBC Count (x 10<sup>9</sup>/L)<sup>‡</sup> | 4326, 0.1±1.9 | 1387, -0.3±1.9 |
| % of Neutrophils<sup>†</sup> | 3312, 66.9±9.9 | 1184, 67.0±10.4 |
| Change in % of Neutrophils<sup>‡</sup> | 3026, 0.4±7.7 | 1102, 0.5±8.4 |
| % of Lymphocytes<sup>†</sup> | 3312, 19.3±8.2 | 1184, 19.4±8.6 |
| Change in % of Lymphocytes<sup>‡</sup> | 3026, -0.2±5.7 | 1102, -0.4±6.3 |
| % of Monocytes<sup>†</sup> | 3312, 6.5±2.0 | 1184, 6.8±2.2 |
| Change in % of Monocytes<sup>‡</sup> | 3026, 0.03±1.7 | 1102, 0.3±2.1 |
| % of Eosinophils<sup>†</sup> | 3312, 4.5±3.0 | 1183, 3.9±3.0 |
| Change in % of Eosinophils<sup>‡</sup> | 3024, -0.2±2.3 | 1101, -0.4±2.6 |
| % of Basophils<sup>†</sup> | 3310, 0.7±0.5 | 1183, 0.6±0.4 |
| Change in % of Basophils<sup>‡</sup> | 3019, -0.02±0.6 | 1099, -0.0003±0.5 |
| NLR<sup>†</sup> | 3312, 4.6±4.3 | 1184, 4.7±3.7 |
| Change in NLR<sup>‡</sup> | 3026, 0.1±4.5 | 1102, 0.4±2.8 |
| MLR<sup>†</sup> | 3312, 0.4±0.3 | 1184, 0.4±0.3 |
| Change in MLR<sup>‡</sup> | 3026, 0.01±0.2 | 1102, 0.04±0.2 |

All variables were included in the ML prediction model to classify the risk of an individual HD patient having a SARS-CoV-2 infection being identified in the following $\geq 3$ days.
† Mean values of laboratory variables 1-14 days before the prediction date (i.e. 3 days before suspicion of SARS-CoV-2 infection in standard clinical practice).
‡ Mean values of the difference in laboratory variables 31-60 days to 1-14 days before the prediction date.
HD: hemodialysis; Hgb: hemoglobin; WBC: white blood cell; TSAT: transferrin saturation; URR: urea reduction ratio; iPTH: intact parathyroid hormone; BUN: blood urea nitrogen; NLR: neutrophil to lymphocyte ratio; MLR: monocyte to lymphocyte ratio; N: patient count; SD: standard deviation.

### Statistical Methods

#### Descriptive Statistics

Descriptive statistics were tabulated for demographics and variables before the predicted COVID-19 outbreak in a county or HD patient SARS-CoV-2 infection.

#### Analysis of ML Model Feature Importance

Shapley values (27, 28) were calculated using the SHAP python package to determine the influence of each variable on model predictions (29, 30). SHAP value is calculated for each variable and each observation, representing a measure of impact (positive or negative) of the observed value’s contribution to each individual prediction. Overall feature importances for each model were calculated using the mean absolute values for each variable across all measures.

#### Analysis of ML Model Performance

Performance of ML models was measured by the area under the receiver operating characteristic curve (AUROC) in the training, validation, and testing datasets, as well as, the recall, precision, and lift in the testing datasets (Refer to **Supplementary Methods** for further details). AUROC, recall, and precision metrics yield scores on a scale of 0 (lowest) to 1 (highest). Lift metric yields an estimate of how many times more/less effective the model is at predicting the outcome versus not using a model. Cutoff thresholds for classifying predictions were selected to optimize recall, precision, and lift.

## Results

### Prediction of Counties with a COVID-19 Outbreak

#### County Characteristics

Among all counties with dialysis clinic(s) servicing local HD populations, 4.9% (n=82) of counties had a major COVID-19 outbreak during the week of 22-28 Mar 2020 (**Supplementary Table 1**), as defined as being in the top 300 United States counties with the highest growth in COVID-19+ people (number and ppm) in the general population (**Table 1**). Counties with a COVID-19 outbreak had a 30% higher population density compared to counties without an outbreak.

On average, the proportion of patients residing at an assisted living facility did not remarkably differ for local HD populations among counties with a major COVID-19 outbreak or not (**Table 1**). The proportion of HD treatments with high body temperatures (>100 F) in the 14 days before a COVID-19 outbreak was <1 percentage point different between county groups. The percent change in the 14-day county-wide mean value for clinical, treatment, and laboratory parameters during the prior 28 days before a COVID outbreak was small in local HD populations and unremarkable in all cases.

#### County-Level Prediction Model Feature Importance

Assessment of variable feature importance with SHAP values showed the top predictors of counties experiencing an outbreak were the COVID-19+ ppm in the general population during prior 7 days, number of deaths per COVID-19+ case in the general population during prior 7 days, and the change in 14-day mean pre-HD body temperature among local HD patients in prior 28 days (**Figure 2A**).

**Figure 2:**
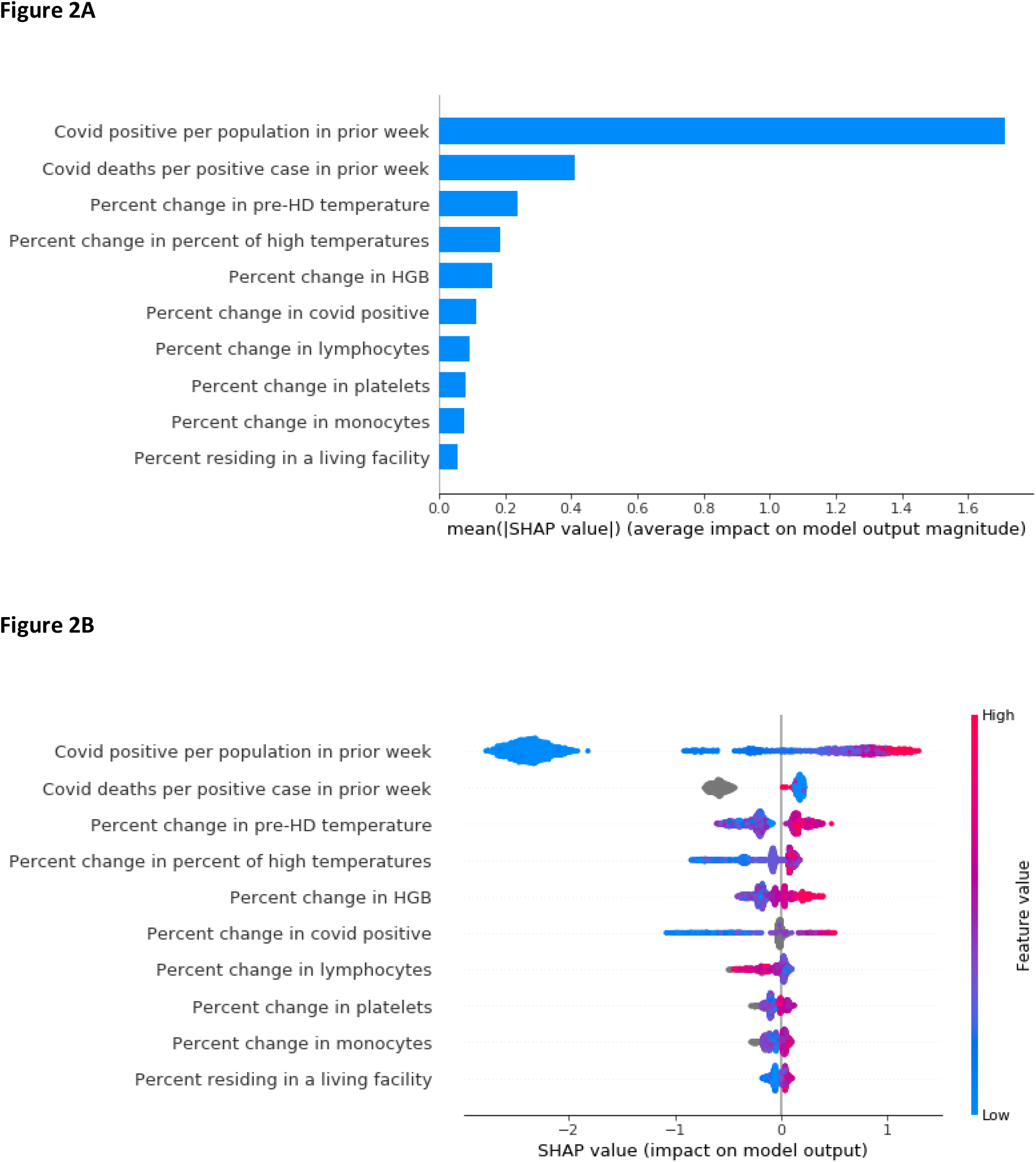
SHAP value plots for the county-level COVID-19 outbreak ML model showing the extent each predictor contributes (positively or negatively) to each individual prediction. **A)** Bar plot of the mean absolute SHAP values for the top 10 predictors in descending order. **B)** SHAP value plot for the degree of the positive or negative impact of each individual measurement on the prediction (x-axis), with warmer colors representing higher observed values for that measurement, and cooler colors indicating lower values for that measurement.

SHAP value plot in **Figure 2B** further shows the degree of positive or negative impact of each individual measurement on the prediction. For example, individual counties with a higher number of COVID-19+ ppm in the general population during prior 7 days (warmer colors) were associated with a higher SHAP value, and this was typically vice versa for individual counties with a lower number of COVID-19+ ppm (cooler colors).

#### County-Level Prediction Model Performance

The ML model had good performance in prediction of the 7-day risk for counties experiencing a COVID-19 outbreak. The AUROC for the model was 0.988, 0.799, and 0.857 in the training, validation, and testing datasets respectively (**Figure 3**).

**Figure 3:**
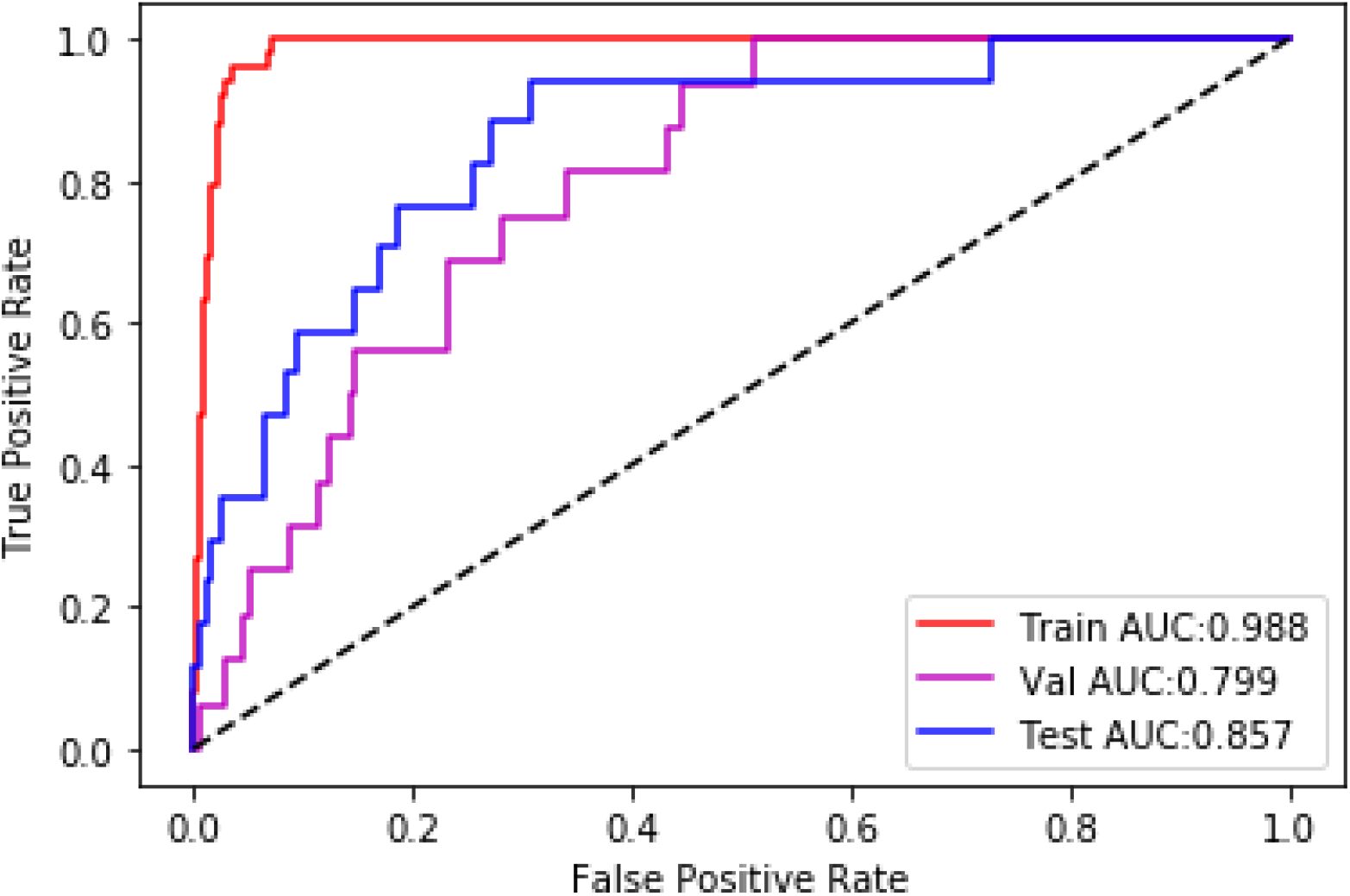
Area under the receiver operating characteristic curve (AUROC) plot for the county-level COVID-19 outbreak ML model showing the rate of true and false positives classified by the prediction model across probability thresholds.

A threshold of 0.85 provided the best performance for the model within goals of limiting false positives. In the testing dataset, the recall was 0.35 showing the model correctly predicted true positives for a COVID-19 outbreak in 35% of positive counties. The precision was 0.40 showing 40% of counties predicted to have an outbreak physically experienced a COVID-19 outbreak. The lift was 7.9 in the testing dataset, suggesting model use is 7.9 times more effective in predicting a COVID-19 outbreak in a county compared to not using a model.

### Prediction of HD Patients with a SARS-CoV-2 Infection

#### Patient Characteristics

We identified data from 11,664 HD patients meeting eligibility criteria (3,249 COVID-19+ cases and 8,415 unaffected (control) patients). The prevalence of COVID-19+ cases (about 28% COVID-19+) in the training and validation datasets was consistent within the cohort. For the testing dataset used to evaluate final model performance, there was a 10% prevalence of COVID-19+ cases based on the designed data split.

In the cohort, there was a higher proportion of HD patients with a SARS-CoV-2 infection of black race, Hispanic ethnicity, and with diabetes (**Table 2**). Mean values for treatment and laboratory variables before a SARS-CoV-2 infection being identified in the subsequent ≥3 days (or concurrent index date in controls) are shown in **Tables 3 & 4**.

**Table 2:** Demographics and Comorbidities of HD Patients with and without SARS-CoV-2 Infection Identified in the Subsequent ≥3 Days

| Variable | Unaffected Patients<br>N (%) or Mean±SD | COVID-19+ Patients<br>N (%) or Mean±SD |
| --- | --- | --- |
| Number of HD patients | 8,415 | 3,249 |
| Age (Years) | 62.75±14.08 | 64.22±13.82 |
| Male | 4694 (55.78%) | 1878 (57.80%) |
| White Race | 3803 (45.19%) | 1219 (37.52%) |
| Black Race | 2158 (25.64%) | 1118 (34.41%) |
| Other Race | 235 (2.79%) | 97 (2.99%) |
| Unknown Race | 2219 (26.37%) | 815 (25.08%) |
| Hispanic Ethnicity | 562 (9.43%) | 402 (16.81%) |
| BMI (kg/m<sup>2</sup>) | 29.60±7.78 | 29.25±8.10 |
| Dialysis Vintage (Years) | 3.66±4.01 | 3.97±4.05 |
| Diabetes | 5588 (66.43%) | 2336 (72.21%) |
| CHF | 2075 (24.67%) | 805 (24.88%) |
| Ischemic heart disease | 2378 (28.27%) | 810 (25.04%) |
| HD: hemodialysis; CHF: congestive heart failure; BMI: body mass index; N: patient count; SD: standard deviation |  |  |

HD patients who contracted COVID-19 had only subtle, clinically unremarkable distinctions in treatment/laboratory characteristics before being suspected to have a SARS-CoV-2 infection compared to unaffected patients. Mean pre-/post-HD body temperatures (**Table 3**) and inflammatory markers (white blood cell (WBC) count and differential) (**Table 4**) before a SARS-CoV-2 infection being identified did not did not show a clinically relevant difference differ between groups. HD patients who had a SARS-CoV-2 infection identified in the following 3 days did have higher ferritin levels compared to unaffected patients.

#### Patient-Level Prediction Model Feature Importance

Calculation of variable feature importance with SHAP values found the top three predictors of HD patients having a SARS-CoV-2 infection were the patient’s mean pre-HD body temperature, blood urea nitrogen (BUN), and albumin (**Figure 4A**).

**Figure 4:**
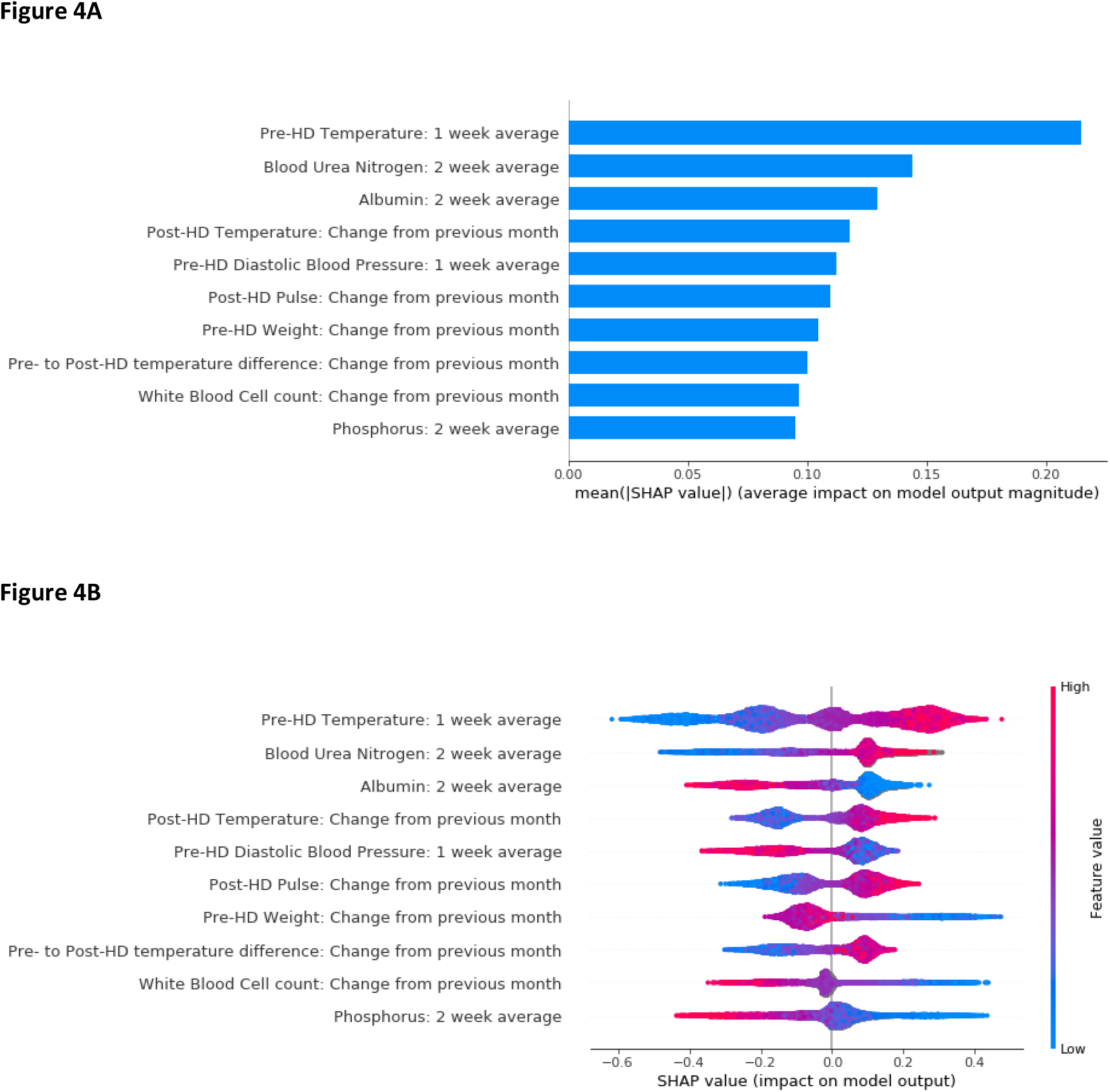
SHAP value plots for the patient-level SARS-CoV-2 infection ML model showing the extent each predictor contributes (positively or negatively) to each individual prediction. **A)** Bar plot of the mean absolute SHAP values for the top 10 predictors in descending order. **B)** SHAP value plot for the degree of the positive or negative impact of each individual measurement on the prediction (x-axis), with warmer colors representing higher observed values for that measurement, and cooler colors indicating lower values for that measurement.

The SHAP value plot is shown in (**Figure 4B**). For the top predictor of mean pre-HD body temperature before a SARS-CoV-2 infection, higher body temperatures (warmer colors) were mostly associated with a positive SHAP value, while lower body temperatures (cooler colors) were always associated with a negative SHAP value.

#### Patient-Level Prediction Model Performance

The HD patient-level model had adequate performance in prediction of the 3-day risk of a SARS-CoV-2 infection. The ML model had an AUROC of 0.88, 0.69, and 0.71 in the training, validation, and testing datasets respectively (**Figure 5**).

**Figure 5:**
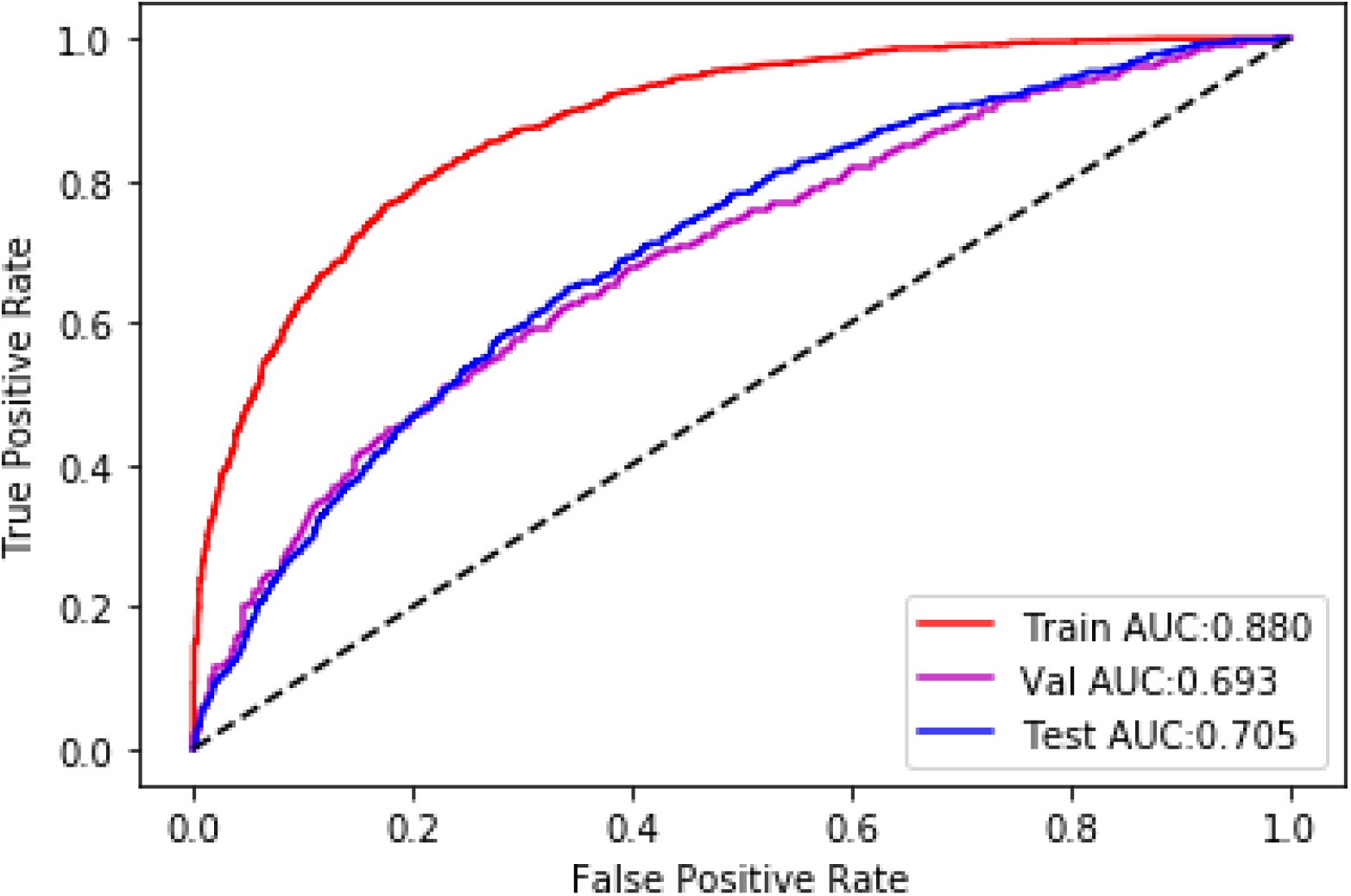
Area under the receiver operating characteristic curve (AUROC) plot for the patient-level SARS-CoV-2 infection ML model showing the rate of true and false positives classified by the prediction model across probability thresholds.

Setting the threshold for classifying observations as positive or negative at 0.85 to minimize false positives, precision for the ML model in the testing dataset was 0.42 showing 42% of patients predicted to have a SARS-CoV-2 infection actually had symptoms in the subsequent ≥3 days and were confirmed to have COVID-19. The lift was 4.2, suggesting model use is 4.2 times more effective in predicting a HD patient who contracts COVID-19, as compared to not having a model. However, given the high threshold, recall was 0.04 showing the model correctly predicted true positives for a SARS-CoV-2 infection in 4% of positive HD patients.

## Discussion

We successfully developed two ML prediction models using retrospective data that appear to have suitable performance in identifying counties at risk of a major COVID-19 outbreak in the following week and HD patients at risk of having a SARS-CoV-2 infection in the following ≥3 days. The top predictor of a COVID-19 outbreak in a county was the COVID-19+ ppm in the general population during prior week. The top predictor of a SARS-CoV-2 infection was the individual patient’s mean weekly body temperature.

Albeit top predictors are not surprising, the observed distinctions were subtle. Without insights from the models considering an array of variables, it would not be clear where one should classify a higher/lower risk for a county or patient that is meaningful. For instance, an increase of 0.2°F (0.1°C) observed in weekly pre-HD body temperature alone may not be considered actionable. The average pre-HD body temperature was 97.5°F (36.4°C) (primarily oral measurements) in our analysis and has been previously reported as 98.2°F (36.7°C) (31). Given 98.6°F (37°C) is the expected average in healthy populations, the lower body temperature of HD patients is of importance with the rather low incidence of fever presenting in dialysis patients with COVID-19 (11%-to-66% with fever (15, 17)).

Prospective testing appears warranted and we anticipate combination of the county- and patient-level models may yield the greatest early insights on where and when providers can focus additional resource allocations to combat COVID-19, and what otherwise asymptomatic HD patients might be most appropriate for COVID-19 testing and triage to an isolation shift/clinic. These models have potential to provide a data-driven way for dialysis providers/clinicians to predict local transmission rates of COVID-19 and individuals with undetected infections. As more data is captured in the COVID-19 outbreak, further prediction models that can classify the risk of morbid/mortal outcomes in dialysis patients need to be developed.

The potential applications of AI for COVID-19 have been previously detailed (32); the first priority was suggested as “early detection and diagnosis of the infection”. In the United States, county-level surveillance models have been developed to estimate dynamics of COVID-19 on hospital resource capacities (33) and relative risk of an emerging COVID-19 outbreak (34). Our model expands upon surveillance efforts by using advanced ML techniques to identify the risks of a COVID-19 outbreak in counties considering predictor variables specific to the HD population. The robustness of data and an *a priori* selection of variables to be included in both the county- and patient-level ML models bring value through assessment of feature importance; this allows for interpretation of meaningfulness of predictors, albeit it does not determine causality.

A systematic review identified several models developed using data from China for early detection of COVID-19 in suspected individuals (23). None were designed for chronic disease populations. One is an externally validated ML model that predicts COVID-19 in suspected asymptomatic patients (AUROC validation=0.872) (35). Another effort used a prediction model (AUROC validation=0.966) to develop logic for an 8 variable COVID-19 risk chart (36). A further model with an AUROC of 0.938 was created to detect COVID-19 pneumonia in patients admitting to a fever clinic (37). Other models use genomic/computed tomography data to diagnose COVID-19 (23). Consistent variables used across models included age, body temperature, and flu-like illness symptoms (23). Although these models were all reported to have suitable performance, all were subject to bias due to non-generalizable sampling of controls without COVID-19 and possible overfitting. We cannot rule out that our ML models may have similar bias, although they included large samples and the testing dataset for the patient-level model had relatively generalizable sampling for the dialysis population with respect to positives/negatives (13). Our patient-level model is unique in its ability to identify the risk of SARS-CoV-2 infection in patients without any suspicion of being affected with the disease.

These developed models hold promise to help providers with the COVID-19 pandemic and any subsequent wave(s) of outbreak (38, 39). Nonetheless, AI modeling is never 100% accurate and model risk classifications need to be interpreted within the extent of the model’s performance.

## Conclusions

The developed AI models showed suitable performance in prediction of dialysis service areas at risk of becoming a COVID-19 hotspot and individual HD patients at risk of having an undetected SARS-CoV-2 infection. Prospective testing is needed, and these models should provide key insights for consideration by healthcare providers.

## Data Availability

The datasets with dialysis patient data used for this analysis are not publicly available.

## Disclosures

CM, JWL, SC, HH, YJ, LAU, FWM are employees of Fresenius Medical Care in the Global Medical Office. KMB, EDW, IADS, KB, JLH, RJK are employees of Fresenius Medical Care North America. LN is an employee of Fresenius Medical Care Deutschland GmbH in the EMEA Medical Office. PK is an employee of Renal Research Institute, a wholly owned subsidiary of Fresenius Medical Care. IADS, KB, PK, JLH, RJK, LAU, FWM have share options/ownership in Fresenius Medical Care. PK receives honorarium from Up-To-Date and is on the Editorial Board of Blood Purification and Kidney and Blood Pressure Research. JLH has directorships in the Renal Physicians Association Board of Directors and Nephroceuticals LLC Scientific Advisory Board. FWM has directorships in the Fresenius Medical Care Management Board, Goldfinch Bio, and Vifor Fresenius Medical Care Renal Pharma. JPK has no conflicts of interest to disclose.

## Authors’ contributions

Design was performed by: CM, IADS, KB, PK, JLH, RJK, LAU, FWM. Data collection and analysis was performed by: CM, JWL, SC, HH, YJ, LAU. The interpretation, drafting and revision of this manuscript was conducted by all authors. The decision to submit this manuscript for publication was jointly made by all authors and the manuscript was confirmed to be accurate and approved by all authors.

## Other contributions

We would like to acknowledge Vladimir M Rigodon for assistance with the composition of the regulatory protocol for this analysis.

## Funding

Project and manuscript composition were supported internally by Fresenius Medical Care.

## References

1. Kliger AS, Silberzweig J: Mitigating Risk of COVID-19 in Dialysis Facilities. Clin J Am Soc Nephrol, 2020

2. Ikizler TA: COVID-19 and Dialysis Units: What Do We Know Now and What Should We Do? Am J Kidney Dis, 2020

3. Basile C, Combe C, Pizzarelli F, Covic A, Davenport A, Kanbay M, Kirmizis D, Schneditz D, van der Sande F, Mitra S: Recommendations for the prevention, mitigation and containment of the emerging SARS-CoV-2 (COVID-19) pandemic in haemodialysis centres. Nephrology, dialysis, transplantation : official publication of the European Dialysis and Transplant Association - European Renal Association, 2020

4. Mokrzycki MH, Coco M: Management of hemodialysis patients with suspected or confirmed COVID-19 infection: perspective of two nephrologists in the United States. Kidney 360: 10.34067/KID.0001452020, 2020

5. Gallieni M, Sabiu G, Scorza D: Delivering safe and effective hemodialysis in patients with suspected or confirmed COVID-19 infection: a single-center perspective from Italy. Kidney360: 10.34067/KID.0001782020, 2020

6. United States Renal Data System. 2019 USRDS annual data report: Epidemiology of kidney disease in the United States. National Institutes of Health, National Institute of Diabetes and Digestive and Kidney Diseases, Bethesda, MD, 2019

7. Roncon L, Zuin M, Rigatelli G, Zuliani G: Diabetic patients with COVID-19 infection are at higher risk of ICU admission and poor short-term outcome. J Clin Virol, 127: 104354, 2020

8. Guo T, Fan Y, Chen M, Wu X, Zhang L, He T, Wang H, Wan J, Wang X, Lu Z: Cardiovascular Implications of Fatal Outcomes of Patients With Coronavirus Disease 2019 (COVID-19). JAMA Cardiol, 2020

9. Li X, Xu S, Yu M, Wang K, Tao Y, Zhou Y, Shi J, Zhou M, Wu B, Yang Z, Zhang C, Yue J, Zhang Z, Renz H, Liu X, Xie J, Xie M, Zhao J: Risk factors for severity and mortality in adult COVID-19 inpatients in Wuhan. J Allergy Clin Immunol, 2020

10. Cheng Y, Luo R, Wang K, Zhang M, Wang Z, Dong L, Li J, Yao Y, Ge S, Xu G: Kidney disease is associated with in-hospital death of patients with COVID-19. Kidney international, 2020

11. Du RH, Liang LR, Yang CQ, Wang W, Cao TZ, Li M, Guo GY, D.J, Zheng CL, Zhu Q, Hu M, Li XY, Peng P, Shi HZ: Predictors of Mortality for Patients with COVID-19 Pneumonia Caused by SARS-CoV-2: A Prospective Cohort Study. Eur Respir J, 2020

12. Adams ML, Katz DL, Grandpre J: Population-Based Estimates of Chronic Conditions Affecting Risk for Complications from Coronavirus Disease, United States. Emerg Infect Dis, 26, 2020

13. Neumann ME: Latest data show 305 dialysis patient deaths due to COVID-19 in the US. Nephrology News & Issues, (Accessed 22 Apr 2020): https://www.healio.com/nephrology/infection-control/news/online/%7B3a263aa269-ad259-264c263f-aab267-207b8395508e8395505%8395507D/latest-data-show-8395305-dialysis-patient-deaths-due-to-covid-8395519-in-the-us, 2020

14. Team CC-R: Geographic Differences in COVID-19 Cases, Deaths, and Incidence - United States, February 12-April 7, 2020. MMWR Morb Mortal Wkly Rep, 69: 465–471, 2020

15. ERACODA - The ERA-EDTA COVID-19 Database for Patients on Kidney Replacement Therapy. ERA-EDTA, (Accessed 02 May 2020): https://www.era-edta.org/en/wp-content/uploads/2020/2004/ERACODA-Study-Report-2020-2004-2029.pdf, 2020

16. Wang H: Maintenance Hemodialysis and Coronavirus Disease 2019 (COVID-19): Saving Lives With Caution, Care, and Courage. Kidney Med, 2020

17. Ma Y, Diao B, Lv X, Zhu J, Liang W, Liu L, Bu W, Cheng H, Zhang S, Yang L, Shi M, Ding G, Shen B, Wang H: 2019 novel coronavirus disease in hemodialysis (HD) patients: Report from one HD center in Wuhan, China. medRxiv: 2020.2002.2024.20027201, 2020

18. Siordia JA, Jr.: Epidemiology and clinical features of COVID-19: A review of current literature. J Clin Virol, 127: 104357, 2020

19. Niiler E: An AI Epidemiologist Sent the First Warnings of the Wuhan Virus. Wired, (Accessed 22 Apr 2020): https://www.wired.com/story/ai-epidemiologist-wuhan-public-health-warnings/, 2020

20. Bogoch, II, Watts A, Thomas-Bachli A, Huber C, Kraemer MUG, Khan K: Pneumonia of unknown aetiology in Wuhan, China: potential for international spread via commercial air travel. J Travel Med, 27, 2020

21. McCall B: COVID-19 and artificial intelligence: protecting health-care workers and curbing the spread. Lancet Digit Health, 2: e166–e167, 2020

22. Alimadadi A, Aryal S, Manandhar I, Munroe PB, Joe B, Cheng X: Artificial intelligence and machine learning to fight COVID-19. Physiol Genomics, 52: 200–202, 2020

23. Wynants L, Van Calster B, Bonten MMJ, Collins GS, Debray TPA, De Vos M, Haller MC, Heinze G, Moons KGM, Riley RD, Schuit E, Smits LJM, Snell KIE, Steyerberg EW, Wallisch C, van Smeden M: Prediction models for diagnosis and prognosis of covid-19 infection: systematic review and critical appraisal. BMJ, 369: m1328, 2020

24. Smith M, Yourish K, Almukhtar S, Collins K, Ivory D, Harmon A, et al.: Coronavirus (Covid-19) Data in the United States: Data from The New York Times, based on reports from state and local health agencies. The New York Times, (Accessed 04 May 2020): https://github.com/nytimes/covid-19-data,

25. Health Resources & Services Administration: Area Health Resources Files. US Department of Health & Human Services, (Accessed 04 May 2020): https://data.hrsa.gov/topics/health-workforce/ahrf,

26. Chen T, Guestrin C: XGBoost: A Scalable Tree Boosting System. Proceedings of the 22nd ACM SIGKDD International Conference on Knowledge Discovery and Data Mining. San Francisco, California, USA, Association for Computing Machinery, 2016 pp 785–794

27. Shapley LS: “A Value for n-Person Games,” In: H. W. Kuhn and A. W. Tucker, Eds., Contributions to the Theory of Games II. Annals of Mathematics Studies, Princeton University Press, Princeton, 28: 307–317, 1953

28. Štrumbelj E, Kononenko I: Explaining prediction models and individual predictions with feature contributions. J Knowledge and Information Systems, 41: 647–665, 2013

29. Lundberg S, Lee SI: “A Unified Approach to Interpreting Model Predictions.” In: I. Guyon, U. V. Luxburg, S. Bengio, H. Wallach, R. Fergus, S. Vishwanathan and R. Garnett, Eds., Advances in Neural Information Processing Systems 30. Curran Associates, Inc: 4765-4774, 2017

30. Lundberg SM, Erion G, Chen H, DeGrave A, Prutkin JM, Nair B, Katz R, Himmelfarb J, Bansal N, Lee S-I: From local explanations to global understanding with explainable AI for trees. Nature Machine Intelligence, 2: 56–67, 2020

31. Usvyat LA, Kotanko P, van der Sande FM, Kooman JP, Carter M, Leunissen KM, Levin NW: Circadian variations in body temperature during dialysis. Nephrology, dialysis, transplantation : official publication of the European Dialysis and Transplant Association - European Renal Association, 27: 1139–1144, 2012

32. Vaishya R, Javaid M, Khan IH, Haleem A: Artificial Intelligence (AI) applications for COVID-19 pandemic. Diabetes Metab Syndr, 14: 337–339, 2020

33. Branas CC, Rundle A, Pei S, Yang W, Carr BG, Sims S, Zebrowski A, Doorley R, Schluger N, Quinn JW, Shaman J: Flattening the curve before it flattens us: hospital critical care capacity limits and mortality from novel coronavirus (SARS-CoV2) cases in US counties. medRxiv: 2020.2004.2001.20049759, 2020

34. Desjardins MR, Hohl A, Delmelle EM: Rapid surveillance of COVID-19 in the United States using a prospective space-time scan statistic: Detecting and evaluating emerging clusters. Appl Geogr, 118: 102202, 2020

35. Meng Z, Wang M, Song H, Guo S, Zhou Y, Li W, Zhou Y, Li M, Song X, Zhou Y, Li Q, Lu X, Ying B: Development and utilization of an intelligent application for aiding COVID-19 diagnosis. medRxiv: 2020.2003.2018.20035816, 2020

36. Song C-Y, Xu J, He J-Q, Lu Y-Q: COVID-19 early warning score: a multi-parameter screening tool to identify highly suspected patients. medRxiv: 2020.2003.2005.20031906, 2020

37. Feng C, Huang Z, Wang L, Chen X, Zhai Y, Zhu F, Chen H, Wang Y, Su X, Huang S, Tian L, Zhu W, Sun W, Zhang L, Han Q, Zhang J, Pan F, Chen L, Zhu Z, Xiao H, Liu Y, Liu G, Chen W, Li T: A Novel Triage Tool of Artificial Intelligence Assisted Diagnosis Aid System for Suspected COVID-19 pneumonia In Fever Clinics. medRxiv: 2020.2003.2019.20039099, 2020

38. Leung K, Wu JT, Liu D, Leung GM: First-wave COVID-19 transmissibility and severity in China outside Hubei after control measures, and second-wave scenario planning: a modelling impact assessment. Lancet, 2020

39. Xu S, Li Y: Beware of the second wave of COVID-19. Lancet, 2020

